# Dietary Iodine Intake in Urban and Rural North India: Implications for and Compatibility with Salt Reduction Efforts

**DOI:** 10.1101/2025.03.10.25323709

**Authors:** Sailesh Mohan, Aprajita Kaushik, Priti Gupta, Kalpana Singh, Ruby Gupta, Dorairaj Prabhakaran

**Affiliations:** Public Health Foundation of India; Centre for Chronic Conditions and Injuries (CCCI), Deputy Director and Secretary,CCDC, Plot 47, Sector 44, NCR, Gurgaon, Haryana, India-122002; Saw Swee Hock School Of Public Health, National University of Singapore, MD 1 Tahir Foundation Building, 12 Science Drive 2, Singapore. 117549; Centre for Chronic Disease Control, Research department, C-1/52, 2nd Floor, Safdarjung Development Area, New Delhi-110016, India; Hamad medical corporation Qatar; Centre for Chronic Disease Control, C-1/52, 2nd Floor, Safdarjung Development Area, New Delhi-110016, India; Public Health Foundation of India (PHFI), Plot 47, Sector 44, NCR, Gurgaon, Haryana, India-122002; Centre for Chronic Disease Control (CCDC), C1/52, 2nd Floor, Safdarjung Development Area, New Delhi, India – 110016

## Abstract

**Objective:** The World Health Organization (WHO) recommends reducing dietary salt intake to <5gm/d as a cost-effective strategy to reduce hypertension and associated cardiovascular diseases. However, salt is also used as a vehicle for iodine supplementation for preventing iodine deficiency disorders (IDD). Concerns have been expressed that reducing population salt intake will adversely impact iodine levels. Therefore, the aim of this paper was to estimate the daily salt and iodine consumption levels of the adult population in India and examine the effect of the WHO recommended salt intake levels on iodine levels.

**Methods:** A cross-sectional study was conducted in Delhi and Haryana, India among adults aged ≥20 years and 24-hr urine samples were collected to assess the salt and iodine levels .

**Findings:** The mean salt intake was 8.07 gm/d (95% CI: 7.03-9.11), mean urinary iodine concentration (UIC) was 208.90 µg/L (184.53-233.27), and the mean urinary iodine excretion (UIE) was 276.15 µg/24hr (239.68-312.62). Analysis to assess iodine intake adequacy by salt intake levels indicated that the mean UIC [180.35 µg/L (159.49-201.20), as well as UIE [153.67 µg/24hr (127.51-179.83)], were adequate, when salt intake was as per the WHO recommended level i.e., ≤5gm/d.

**Conclusion:** Even at the WHO recommended salt intake levels, population iodine intake in this study population was adequate, likely indicating that upward titration in iodine concentration in salt may not be required. Thus, current salt reduction recommendations are unlikely to impact iodine supplementation efforts, indicating compatibility between the two interventions, which should be continiously monitored through robust surveillance.

## Introduction

The recent global burden of disease (GBD) data reported that in India, noncommunicable diseases (NCDs) are responsible for almost 73.4% of all deaths(1), with cardiovascular diseases (CVDs) accounting for 27.4% and 13.9% of Disability Adjusted Life Years (DALYs) lost. (2)(3)

Excess dietary salt intake is a major risk factor for high blood pressure and CVDs. Thus, salt reduction is recomended by most global health organizations as a key strategy to reduce CVDs. Reducing the salt or sodium intake by 30% by 2025 is among the WHO’s nine voluntary global targets. (3) The most recent WHO status report shows that world is far behind in achieving this target. (4)While the WHO recommends reducing salt intake to less than 5 gm/day (equivalent to 1 teaspoon) per person, it is important that before a salt reduction policy is implemented, the global public health challenge of iodine deficiency disorders (IDD) is also addressed concurrently. (5) As consuming iodised salt is one of the best, least expensive and most effective interventions for preventing Iodine deficiency disorders (IDD), it is essential to synergise implementation of both efforts or programmes. (6)

Most experts agree that the two programmes are compatible and can be further strengthening by an integrated approach to salt reduction and salt iodisation efforts i.e., if a reduction in population salt intake occurs, an upward titration of salt iodine levels can be done with national level monitoring or surveillance to the population intake levels of both. Currently the iodine fortification is done based on the assumption that 10 gm/d of salt intake per person can provide adequate intake of iodine. (6)

However, to achieve this harmonization, accurate data on mean population salt intake and iodine intake levels are required. This data is currently unavailable in India, where the National Multisectoral Action Plan for Prevention and Control of common NCDs is being implemented with salt reduction as one of the components. Therefore, to fill this important evidence gap, a population survey to assess the mean daily dietary salt and iodine intake was conducted among adults residing in urban and rural areas of India. We also report on the impact of the WHO recommended salt intake on iodine intake levels.

## Methods

### Study design

The detailed methods of this cross-sectional survey have been described previously. (7) Briefly, the study sites were selected to include slum and non-slum urban and rural areas. The urban part of the survey was conducted in Delhi and the rural part in Faridabad, Haryana. Participant recruitment was done using a stratified random sampling method to recruit individuals from urban, urban slum, and rural areas, from six age and sex groups. Census enumeration blocks (CEBs) and villages were sampled randomly from within the study area. Households were then selected randomly and one individual from within each household was selected randomly until recruitment members in each stratum were fulfilled. In urban areas, two individuals—one male and one female were selected from a household. The study protocol was approved by the Indian Health Ministry’s Screening Committee (HMSC) and the ethics committee of the Centre for Chronic Disease Control, New Delhi. Written informed consent was obtained from all the participants. (8)

### Data collection

Data was collected by trained field staff by conducting a face-to-face interview at home . Consenting participants provided questionnaire data and underwent measurements of height, weight, waist circumference, and blood pressure. Participants were then provided with detailed verbal and written instructions along with a kit for their 24-hr urine collection. A urine collection day was agreed upon, and a reminder call was placed to the participant the evening before their scheduled starting date.

Weight, height, BMI, and waist circumference were measured using the standard WHO STEPS protocol. (9) Blood pressure was measured using an Omron HEM7121 standard blood pressure monitor. Two measurements were taken with 3-minute intervals between each, and if the second measurement differed by >10 mm Hg for systolic blood pressure or 5 mm Hg for diastolic blood pressure, a third measurement was taken. The average of the 2 measurements, or the last 2 measurements, if a third measurement was taken, was used. (8)

### Determination of urinary iodine

24-Urine samples were collected by trained field staff on the day of completion and transferred to a central laboratory here volume was measured, and an aliquot was drawn for assay. Aliquots were transferred to a central laboratory in Delhi for analysis. Iodine in urine was estimated using the Ammonium Persulfate Digestion on Microplate method. (10) All samples were run in triplicate. Three levels of internal controls were run with every batch of samples. The intra- and inter-assay coefficient of variation was 2.4% and 5.3% respectively. The laboratory successfully participated in the EQUIP program from CDC, Atlanta with a score of 91.6 %. (11)

### Determination of salt iodine

Iodine in salt , which was collected from 205 households, was estimated by iodometric titration with sodium thiosulphate. (12) All samples were run in triplicate with two levels of controls. The intra- and inter-assay coefficient of variation was 1.39% and 1.42% respectively.

### Statistical analyses

Results for urinary iodine concentration are presented as medians with interquartile ranges (IQR) and means with 95% confidence intervals (CI) for all participants who consented to provide a urine sample and had valid urinary iodine results. We accounted for the complex survey design in all analyses by adjusting standard errors for clustering and incorporating sampling weights. Estimates of prevalence and associations were age- and sex-standardized to the 2010 South Asia regional population. Results for salt iodine concentration are presented as means with 95% confidence intervals (CI). According to international guidelines, a median urinary iodine concentration (UIC) below 100 µg/L is insufficient iodine intake; 100-199 µg/L indicates adequate iodine intake, while a median above 199 µg/L indicates iodine intake above requirements. Whereas, a mean urinary iodine excretion (UIE) within the range of 85-600µg/24hr is considered adequate (13). It has been proposed that the estimated UIE is more trustworthy than UIC, since it accounts for variations in urine volume and sample dilution.

For salt, iodine content less than 5 ppm is termed as non-iodised; between 5 and 14.9 ppm as inadequately iodised. If the iodine content is ≥ 15 ppm it is considered as adequately iodised. (14) Multiple linear regression was used to test the relationship of Urinary Iodine Concentration (UIC) (µg/L) and Urinary Iodine Excretion (UIE) (µg/24hr) with demographics, behavioural factors and health related factors. Demographic variables of interest included were; age group (20-44, 45-60, >60); sex (male versus female); occupation (employed versus non-employed), and education (up to primary, secondary, and above secondary). Behavioural factors were: ever tobacco use, ever alcohol use, and physical activity. Health related factors were: blood pressure, history of diabetes, chronic kidney disease (CKD), high cholesterol. The salt-related factors consisted of the type of salt (iodised versus non-iodised), salt intake (≤5gm/d versus >5gm/d); adequacy (ppm), and salt iodine concentration categorised (adequate versus non-adequate).

A p-value of < 0.05 was considered statistically significant. Analyses were performed using Stata software (version 15; Stata Corporation, College Station, TX).

## Results

### Sample characteristics

Of the 1716 persons selected for the survey at the two sites, 1484 agreed for the interview (overall response rate, 86.5%; rural, 76.6%; urban, 97.1 %), and 1208 agreed to provide the 24-urine samples (overall response rate, 70.4%; rural, 71.1%; urban 69.7%). Of the 1208 urine samples, urinary iodine values were available for 1112. The mean age of the participants was 41.7 years (±15.2). The majority of the participants were employed (76.7%), attended secondary school (51.1%), and used iodised salt (99.6%). Approximately, one-third of the participants were tobacco users and hypertensives, while one-fourth ever consumed alcohol **(Table 1)**.

**Table 1:** Characteristics of study participants in Delhi and Haryana (N=1484)

|  | Urban | Urban slum | Rural | Total |
| --- | --- | --- | --- | --- |
| **Mean age in years (SD)** | 45(13.7) | 40(13.7) | 41 (15.6) | 41.7 (15.2) |
| **Age groups (%)** |  |  |  |  |
| 20-45 years | 50.4 [33.7,67.1] | 67.4 [61.6,72.7] | 66.9 [0.2,2.5] | 63.1 [56.9,68.9] |
| 45-60 years | 34.4 [16.9,57.6] | 19.5 [16.5,22.8] | 17.4 [13.8,21.7] | 22.1 [16.3,29.2] |
| >60 years | 15.2 [9.8,22.8] | 13.2 [8.8,19.2] | 15.6 [14.3,17.1] | 14.8 [12.7,17.2] |
| **Gender (%)** |  |  |  |  |
| Female | 54 [40.1,67.4] | 45.9 [41.5,50.3] | 51.3 [44.1,58.4] | 50.4 [45.3,55.6] |
| Male | 46 [32.6,59.9] | 54.1 [49.7,58.5] | 48.7 [41.6,55.9] | 49.6[44.4,54.7] |
| **Highest level of education (%)** |  |  |  |  |
| Primary school | 35.8 [23.0,51.1] | 33.3 [25.7,41.8] | 40.9 [29.7,53.2] | 37.6 [30.4,45.3] |
| Secondary school | 58 [41.5,72.8] | 54.9 [48.0,61.7] | 45.4 [36.5,54.6] | 51.1[44.0,58.1] |
| Above secondary | 6.2 [2.5,14.7] | 11.8 [8.9,15.5] | 13.7 [10.1,18.2] | 11.3 [8.8,14.4] |
| **Employment status (%)** |  |  |  |  |
| Employed /Home maker | 87.4 [77.9,93.2] | 76.9 [65.8,85.2] | 75.6 [73.3,77.7] | 76.7 [74.0,79.2] |
| Unemployed /Student | 12.6 [6.8,22.1] | 23.1 [14.8,34.2] | 24.4 [22.3,26.7] | 23.3[20.8,26.0] |
| **BMI (kg/m<sup>2</sup>) (%)** |  |  |  |  |
| <18 | 5.6 [1.3,21.6] | 10.1 [7.7,13.2] | 12.5 [8.8,17.4] | 10.2 [7.3,14.0] |
| 18-22.9 | 22.1 [12.8,35.3] | 33.1 [27.5,39.2] | 43.6 [40.4,46.8] | 35.5 [30.0,41.4] |
| 23-24.9 | 16.3 [9.8,26.1] | 15.6 [27.5,39.2] | 19 [16.0,22.4] | 17.4[14.2,21.2] |
| >25 | 56 [34.7,75.3] | 41.2 [34.5,48.2] | 24.9 [20.1,30.5] | 36.9 [28.3,46.4] |
| **Tobacco use (%)** |  |  |  |  |
| Yes | 19.2 [9.7,34.4] | 32.9 [28.1,38.1] | 39.4 [33.3,45.7] | 32.7[27.1,38.9] |
| No | 80.8 [65.6,90.3] | 67.1 [61.9,71.9] | 60.6 [54.3,66.7] | 67.3[61.1,72.9] |
| **Ever Alcohol consumption (%)** |  |  |  |  |
| Yes | 20.6 [13.1,30.8] | 20.8 [16.0,26.5] | 21.6 [11.0,38.0] | 21.1[15.0,28.9] |
| No | 79.4 [69.2,86.9] | 79.2 [73.5,84.0] | 78.4 [62.0,89.0] | 78.9 [71.1,85.0] |
| **Physical activity (%)** |  |  |  |  |
| Yes | 96.8 [88.8,99.1] | 93.5 [88.0,96.6] | 95.9 [95.4,96.4] | 95.4 [93.7,96.6] |
| No | 3.2 [0.9,11.2] | 6.5 [3.4,12.0] | 4.1 [3.6,4.6] | 4.6 [3.4,6.3] |
| **Blood pressure (%)** |  |  |  |  |
| Normotensive | 21.5 [12.4,34.6] | 42.6 [37.5,47.8] | 42.7 [39.3,46.3] | 37.6 [32.4,43.1] |
| Pre-hypertensive | 33.7 [22.1,47.6] | 29.5 [25.7,33.7] | 36.7 [32.5,41.1] | 34 [29.9,38.4] |
| Hypertensive | 44.9 [28.3,62.7] | 27.9 [23.8,32.4] | 20.6 [16.7,25.1] | 28.4[21.9,36.1] |
| **Salt usage (%)** |  |  |  |  |
| Iodised | 99.3 [97.5,99.8] | 99.8 [98.2,100.0] | 99.6 [98.8,99.9] | 99.6 [99.1,99.8] |
| Non-iodised | 0.7 [0.2,2.5] | 0.2 [0.0,1.8] | 0.4 [0.1,1.2] | 0.4 [0.2,0.9] |

### Mean salt, UIC (μg/L) and UIE (μg/24hr) levels

Overall, the mean dietary salt intake was 8.07 gm/d (95% CI 7.03-9.11), highest in the urban slum sites (9.41gm/d) **(Table 2, Figure 1)**. The estimated mean UIC was 208.90 µg/L (184.53-233.27), while the mean UIE was 276.15 µg/24hr (239.68-312.62). Both UIC and UIE were higher in rural sites, as compared to the urban and urban slum sites **(Supplementary Table S2, Figure S1)**. UIC indicated adequate iodine intake among all participants from urban [180.22µg/L (202.42-276.23)] and urban slum sites [191.18µg/L (248.11-297.21)] but excess intake for rural sites [299.19µg/L (239.68-312.62)]. However, UIE indicated adequate iodine intake among urban, urban slum, and rural sites.

**Table 2:** Iodine adequacy stratified by demographic characteristics, salt usage, and salt intake (N=1112)

|  | UIC (µg/L) |  | UIE (µg/24hr) |  |
| --- | --- | --- | --- | --- |
|  | Mean | 95% CI | Mean | 95% CI |
| **Highest level of education (%)** |  |  |  |  |
| Primary school | 207.45 | (184.57-230.34) | 270.98 | (226.77-315.18) |
| Secondary school | 210.61 | (179.19-242.03) | 278.6 | (239.86-317.34) |
| Above secondary school | 203.95 | (178.81-229.09) | 273.39 | (232.86-313.93) |
| **Employment status (%)** |  |  |  |  |
| Employed | 227.68 | (197.73-257.62) | 292.25 | (232.65-351.85) |
| Unemployed | 213.43 | (176.39-250.46) | 307.19 | (260.79-353.59) |
| **BMI** |  |  |  |  |
| Below 18 | 199.43 | (161.68-237.18) | 218.65 | (194.26-243.05) |
| 18-22.9 | 210.47 | (173.69-247.25) | 263.38 | (208.16-318.60) |
| 23-24.9 | 208.87 | (163.68-254.07) | 294.37 | (217.79-370.94) |
| >=25 | 208.73 | (177.71-239.74) | 293.46 | (267.54-319.38) |
| **Ever tobacco use** |  |  |  |  |
| Yes | 218.87 | (179.52-258.22) | 283.02 | (227.62-338.42) |
| No | 192.81 | (138.31-247.32) | 235.19 | (169.74-300.64) |
| **Ever alcohol consumption** |  |  |  |  |
| Yes | 230.85 | (198.85-262.84) | 294.22 | (244.9-343.55) |
| No | 202.82 | (179.59-226.05) | 271.29 | (237.14-305.43) |
| **Physical activity** |  |  |  |  |
| Yes | 211.10 | (182.28-239.91) | 282.31 | (239.17-325.45) |
| No | 201.13 | (172.79-229.47) | 255.62 | (197.94-313.30) |
| **Blood pressure** |  |  |  |  |
| Normotension | 206.51 | (178.29-234.73) | 263.97 | (221.01-306.93) |
| Prehypertension | 224.51 | (194.18-254.84) | 283.25 | (238.28-328.22) |
| Hypertension | 193.54 | (158.52-228.56) | 282.5 | (250.07-314.94) |
| **History of stroke** |  |  |  |  |
| Yes | 166.67 | (89.11-244.24) | 213.37 | (150.7-276.04) |
| No | 209.10 | (184.61-233.59) | 276.45 | (239.82-313.08) |
| **History of diabetes** |  |  |  |  |
| Yes | 137.58 | (88.38-186.77) | 268.44 | (230.26-306.61) |
| No | 216.24 | (193.57-238.90) | 277.09 | (238.58-315.60) |
| **History of chronic kidney disease** |  |  |  |  |
| Yes | 39.70 | (22.86-56.55) | 89.93 | (55.1-124.77) |
| No | 223.7 | (193.57-253.83) | 292.9 | (237.76-348.04) |
| **History of high cholesterol** |  |  |  |  |
| Yes | 113.09 | (58.57-167.61) | 262.71 | (243.18-282.24) |
| No | 213.19 | (189.96-236.41) | 276.75 | (238.71-314.79) |
| **Salt usage** |  |  |  |  |
| Iodized | 208.1 | (184.33-231.86) | 273.81 | (238.85-308.77) |
| non-iodized | 179.67 | (35.62-323.71) | 296.41 | (48.25-544.57) |
| **Salt intake** |  |  |  |  |
| ≤5gm/d | 180.35 | (159.49-201.20) | 153.67 | (127.51-179.83) |
| >5gm/d | 222.52 | (190.64-254.41) | 334.61 | (292.26-376.96) |

**Figure 1:**
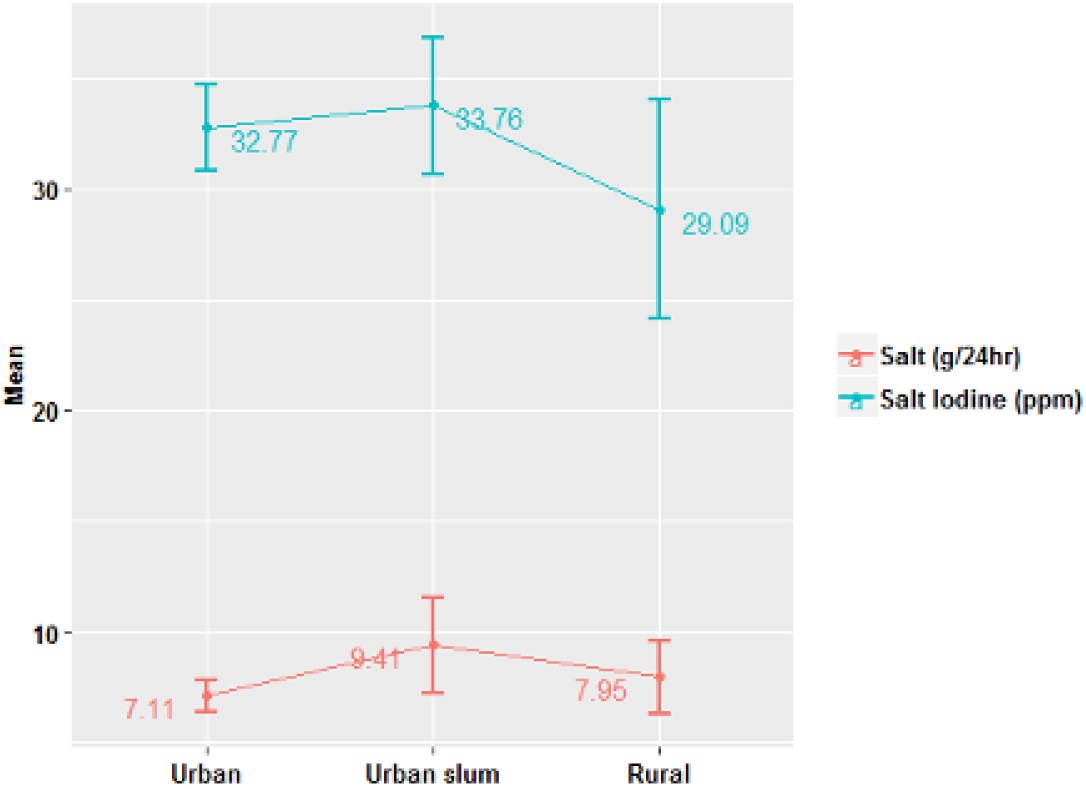
Mean salt intake (gm/d) (N=1112) and mean Iodine intake from household salt (ppm) (N=205) for the urban, urban slum, and rural sites of Delhi and Haryana, India

**Figure 1:** Mean salt intake (gm/d) (N=1112) and mean Iodine intake from household salt (ppm) (N=205) for the urban, urban slum, and rural sites of Delhi and Haryana, India

### SIodine adequacy

#### a. Stratified for demographic, behavioural and health related characteristics

Further analysis to assess iodine adequacy by health related factors indicated excess mean UIC for all, except participants who were hypertensive [193.54 μg/L (158.52-228.56)], had a history of stroke [166.67 μg/L (89.11-244.24)], diabetes [137.58 μg/L (88.38-186.77)] and high cholesterol [113.09 μg/L (58.57-167.61). For participants who reported having chronic kidney disease, iodine intake as indicated by the mean UIC was reported to be inadequate [39.7 μg/L (22.86-56.55)]. For participants who reported using iodised salt, excess iodine intake, as indicated by the mean urinary iodine concentration (UIC), was observed [208.1 µg/L (range: 184.33–231.86)]. However, iodine intake, as measured by the mean urinary iodine excretion (UIE), was found to be adequate across all demographic groups. **(Table 2)**.

#### b. Stratified for salt intake levels

Analysis to assess iodine adequacy by salt intake levels indicated that the mean UIC [180.35 µg/L (159.49-201.20)], as well as UIE, [153.67 µg/24hr (127.51-179.83)], were adequate when the salt intake was ≤5gm/d (15) **(Table 2, Figure 2)**. When salt intake was >5gm/d, the mean UIC was 222.52 µg/L (190.64-254.41). However, UIE was adequate even for participants reporting salt intake >5gm/d **(Table 2, Figure 2)**.

**Figure 2:**
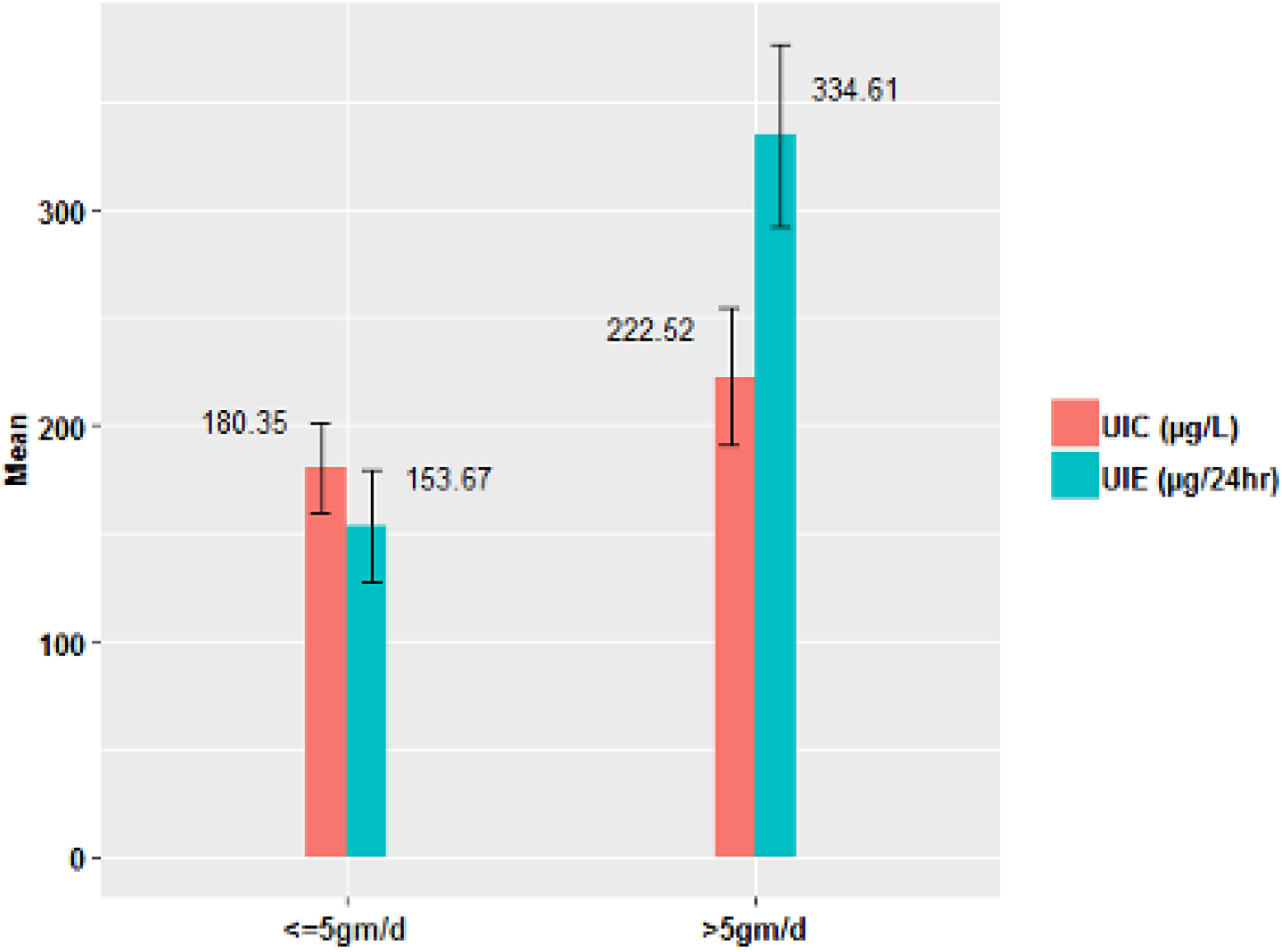
Adequacy of iodine intake by different levels of salt intake (N= 1112)

#### c. Stratified for hypertension

When iodine adequacy was further stratified by salt intake and hypertension status, our analysis indicated that iodine intake by mean UIC was adequate for those who consumed the recommended levels of salt i.e., ≤5gm/d and were normotensive [175.98 µg/L (142.59- 209.36)] or hypertensive [155.37 µg/L (136.18-174.56)]. For participants who consumed >5gm/d salt, iodine intake by mean UIC was in excess, irrespective of their blood pressure status. Iodine intake by mean UIE was adequate for all participants, irrespective of salt intake levels or blood pressure status **(Supplementary table S1)**.

### Mean iodine content (ppm) from household salt

The overall mean iodine content (ppm) from household salt (N=205) was estimated to be 31.66 ppm (29.02-34.31), highest in the urban slum sites [33.76 ppm (30.67-36.85)] , followed by urban sites [32.77 ppm (30.80-34.74)] and rural areas, [29.09 ppm (24.10- 34.08)]**( Figure 1)**. When stratified for demographics such as education, employment, BMI, history of stroke, etc. similar figures for mean iodine content (ppm) were observed

### Multivariate analysis (Table 3) UIC(µg/L)

After adjusting for demographic , behavioural and other health related variables, participants living in rural areas and consuming >5gm/d salt showed a higher salt intake Those who were unemployed showed a lesser salt intake.

**Table 3:**
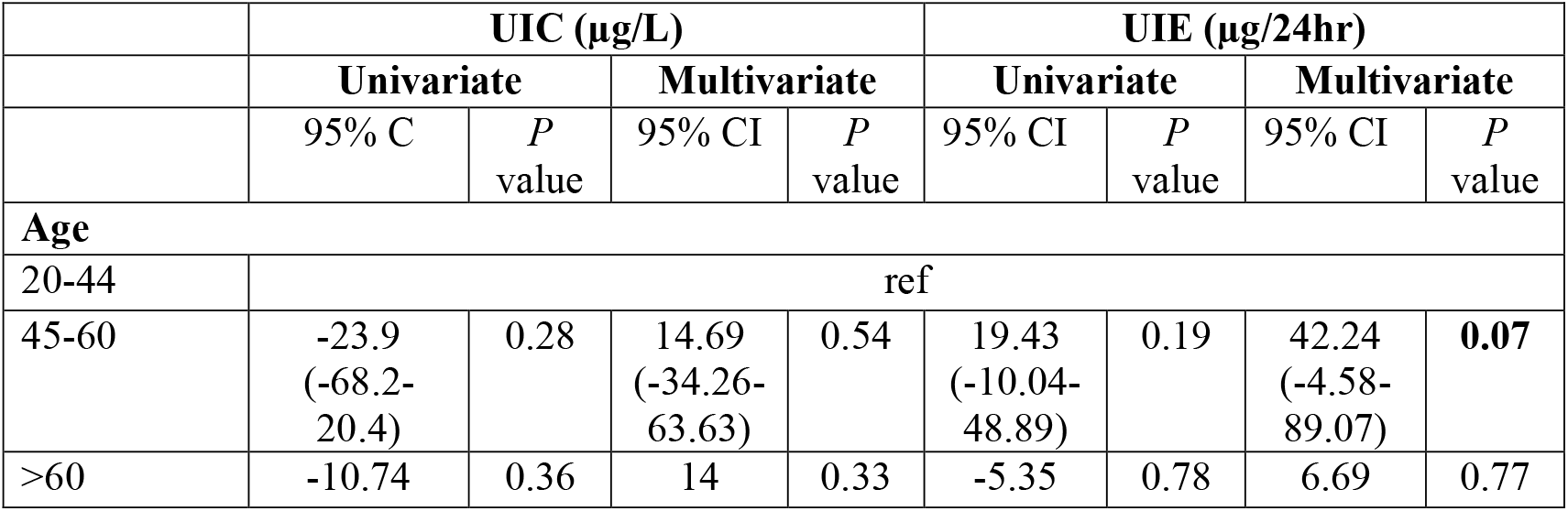
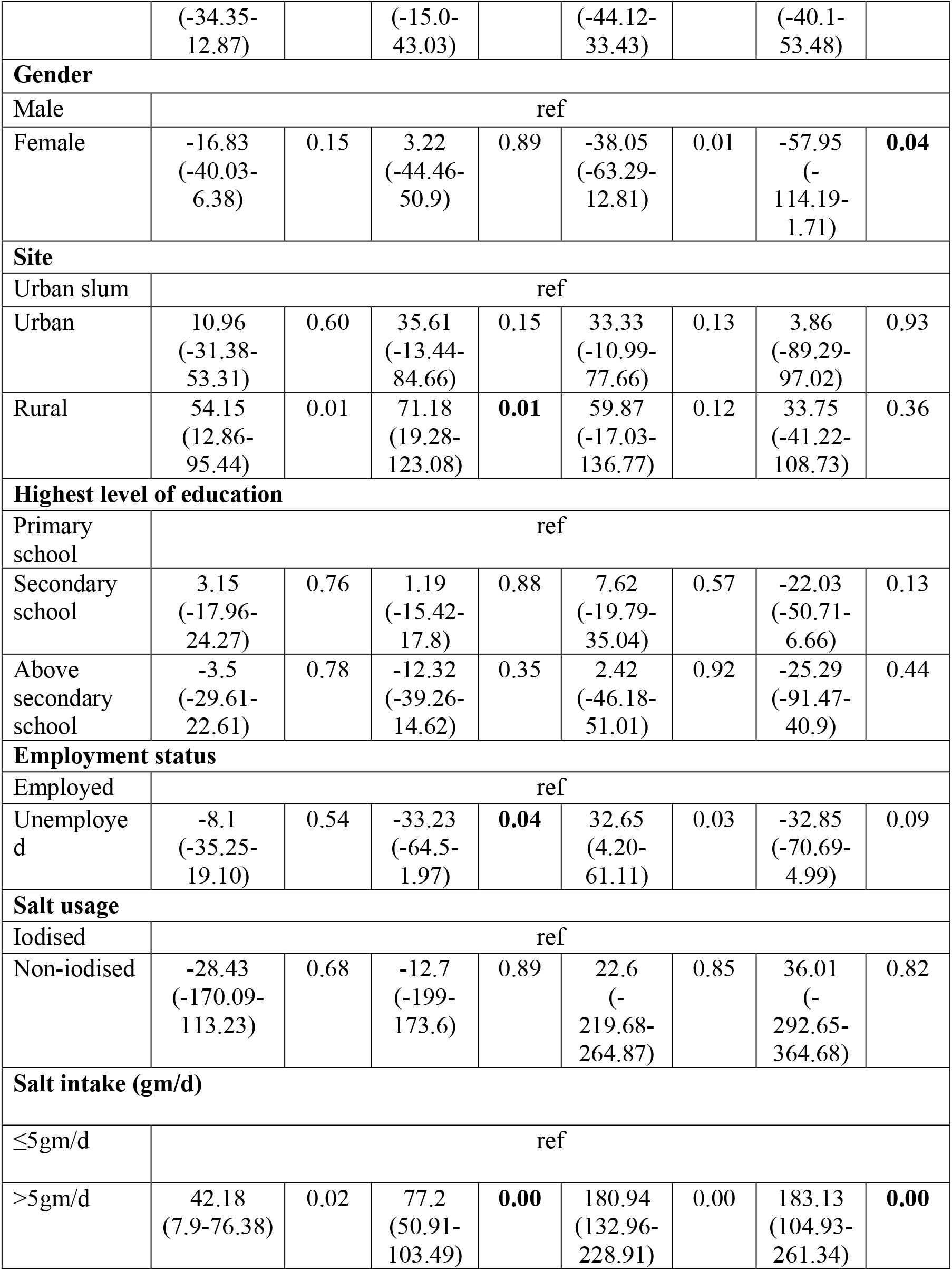
Regression analysis of Urinary Iodine Concentration (UIC) (µg/L) and Urinary Iodine Excretion (UIE) (µg/24hr) estimated from 24-hour urine samples for participants in Delhi and Haryana.

### UIE(µg/24hr)

After adjusting for demographics, behavioural and other health related variables, age (45-60 years), gender and salt intake showed significant associations with UIE (µg/24hr). The age group 45-60 years, showed a positive association [42.24 µg/24hr (95%CI: -4.58-89.07)]. Females showed a negative association [-57.95 µg/24hr (95%CI: -114.19-1.71)], while participants who consumed >5gm/d salt showed a positive association [183.13µg/24hr (104.93-261.34)].

## Discussion

This study indicates that overall mean population dietary salt intake was 8gm/day, which is above the WHO recommended levels of <5gm/day. Both the mean UIC (µg/L), as well as UIE (µg/24hr), were compatible with the recomended salt intake levels i.e., iodine intake was adequate even when salt intake was as per the WHO recommended amount i.e., ≤5gm/d (10). However, UIC (µg/L) was higher when salt intake was >5gm/d. Most of the participants were consuming iodised salt and the mean ppm iodine content was 31.7 ppm.

The iodine intake in our study is similar to the National ‘India Iodine Survey 2018-19’.(16) However, the India iodine survey used only spot urine samples, whereas we used a 24-hr urinary assessment, the Gold standard method to determine iodine levels. Importantly, we also assessed UIE (µg/24hr) along with UIC (µg/L), which provides a better measure of the intake.

Our findings also shed light on most important policy and programmatic concerns surrounding an upward titration of iodine in order to reduce dietary salt intake as per the WHO recommendation (10). Our findings indicate that iodine adequacy at recomended salt intake levels and likely points towards compatibility between the two programmes —salt reduction and salt iodisation. This also indicated that an upward titration of iodine concentartion in salt may not be required in this population. A comparable study undertaken in South Africa explored the compatibility of two programmes, highlighting the necessity of monitoring iodine levels. In this study also urinary iodine excretion (UIE) was aligned with the WHO recommended iodine levels but urinary iodine concentration (UIC) was lower, in individuals consuming less than 5gm/day of salt.(17)

Almost all participants in this study were consuming iodised salt. These findings are in sync with the National Family Health Survey (NFHS-5) 2019-20, where 94.3% of the households (93% in rural and 96.9% in urban) were using iodised salt. (18)

The mean iodine content in salt in our study sample was 31.7ppm, which is well within normal WHO-recommended levels. (19) The recommended minimum level of salt iodine at the household level is 15 ppm in order to achieve the necessary iodine concentration. Perhaps, due to improvements in salt packaging and distribution technology, loss during transportation has been significantly reduced. As a result, the salt iodine content at the household was almost twice as high as the recommended minimum level. Thus, it appears that the present salt iodization levels are sufficient to maintain an adequate iodine level even among those whose daily salt intake was <5gm/day.

## Strengths and limitations

Our study provides hitherto unavailable evidence on the adequacy of iodine intake while advocating for a population salt reduction strategy. This supports the compatibility of implementing programmes to address both iodine deficiency disorders and reducing excess dietary salt intake to prevent and control high blood pressure and CVDs. A salt reduction programme, with an emphasis on consuming iodised salt, will make the two programmes more synergistic, helping us achieve their collective aims.

Most of the previous studies on iodine intake in India were among children and pregnant women and used spot urine samples, but this study was conducted among free-living adults using 24hr urine samples for iodine assessment.

However, this study also has some limitations. Salt iodine levels were based on salt samples collected from the households of 205 participants only. However, we think that this is unlikely to have influenced the results.

## Conclusion

Our study demonstrates that in India where universal salt iodisation is mandatory, a reduction in salt intake to WHO’s recommended levels of ≤5gm/d does not compromise the level of iodine intake among participants as measured by 24-hr urinary iodine concentration (UIC) and urinary iodine excretion (UIE) in adults. These findings provide strong support for the WHO’s recommendations to reduce population salt intake to prevent hypertension and cardiovascular diseases, and also allays concerns regarding the requirement of an upward iodine titration in salt to account for the reduced dietary salt intake. Thus, current salt reduction recommendations are unlikely to impact iodine supplementation efforts, indicating compatibility between the two interventions.

Authors contribution: SM and DP concetulaize the idea, executed the study, reviewed and confirmed the final draft of paper, AK wrote the first draft, PG and KS helped in data analayis and interpretation, RG helped in salt and urine data collection in field and did the laboratory analysis. All authors have read and contributed in paper writing.

## Data Availability

Deidentified Stata files used to conduct the analysis could be provided upon reasonable request made to the corresponding author.

## Acknowledgments

We would like to express our sincere gratitude to our field investigators for their dedicated efforts in collecting both the questionnaires and 24-hour urine samples. We also extend our thanks to our lab team for their thorough analysis of the urine samples. Finally, we are deeply appreciative of our study participants for generously providing both their valuable information and biochemical samples. Gupta Priti is an Early Career Fellow of the WT/DBT India Alliance with grant number IA/CPHE/20/1/505259.

